# Prognostic impact of left ventricular assist device-related complications under the new heart transplant allocation system

**DOI:** 10.64898/2026.01.10.26343868

**Authors:** Timothy Lee, Noah Moss, Nana Toyoda, Natalia Egorova, Gregory Serrao, Mohit Pahuja, Koichi Nomoto, Anelechi C. Anyanwu, Shinobu Itagaki

**Author notes:** **Corresponding author**: Shinobu Itagaki, MD, MSc, Department of Surgery, Division of Cardiothoracic and Vascular Surgery, University of Oklahoma Medical Center, 800 Stanton L Young Blvd, Oklahoma City, OK 73117, USA. This paper was presented at the 104th Annual Meeting of the American Association for Thoracic Surgery, Toronto, Canada, in April 2024.

## Abstract

**Background:** In 2018, the United Network for Organ Sharing (UNOS) revised the donor heart allocation policy, replacing the single urgency status for left ventricular assist device (LVAD)-related complications with three distinct categories. We evaluated the impact of this policy modification on transplant access and outcomes.

**Methods:** The UNOS Standard Transplant Analysis and Research File was queried to identify adult patients listed for heart transplantation with a LVAD-related complication in the United States between 2018 and 2023. The cumulative incidence of heart transplantation, mortality on device, and overall mortality following complication were assessed.

**Results:** During the study period, 792 patients experienced an LVAD complication that led to an initial listing or change in urgency status. Device infection was the most frequent complication (n=472, 59.6%), followed by device malfunction (n=80, 10.1%), aortic regurgitation (n=73, 9.2%), ventricular arrhythmias (n=46, 5.8%), thrombosis/hemolysis (n=43, 5.4%), bleeding (n=42, 5.3%), and right heart failure (n=36, 4.5%). At 1 year, transplantation incidence was 71.5% (95% CI, 67.9–74.8%), mortality on device was 3.8% (95% CI, 2.5–5.4%), and overall mortality was 12.3% (95% CI, 9.9–15.1%). Right heart failure was associated with increased 1-year mortality (34.1%, 95% CI, 18.2–50.8%; adjusted HR 2.0, 95% CI, 1.1–3.8).

**Conclusions:** The revised allocation system provides LVAD patients with complications timely access to transplantation, reflected in high transplant rates and low mortality. Right heart failure remains a distinct subgroup, with one-third of patients not surviving to one year, suggesting this complication may warrant consideration for higher urgency status.

## Introduction

Durable left ventricular assist device (LVAD) is indicated for patients with advanced heart failure to improve functional status, quality of life and survival.^1^ LVAD implantation has become an established therapy for end-stage heart failure, driven both by the shortage of donor hearts and by advances in device technology that have markedly improved survival.^2–4^ In the seminal Randomized Evaluation of Mechanical Assistance for the Treatment of Congestive Heart Failure (REMATCH) trial,^5^ 1-year survival with pulsatile LVADs was 54%; in contrast, contemporary outcomes with the HeartMate 3, the sole implantable LVAD currently available in the United States and Europe, demonstrate 64% survival at 5 years.^4^

Improved on-device survival has the negative consequence of exposing patients on LVADs to a greater amount of time to experience adverse events. This challenge has been highlighted since the 2018 revision of the United Network for Organ Sharing (UNOS) heart allocation system, which assigns stable LVAD patients to Status 4 (of six tiers), a reduction from their prior Status 1B (second-highest of three tiers).^6^ Although newer-generation LVADs have reduced complication rates,^7,8^ the burden of events remains substantial, and allocation policy plays a key role in providing timely access to transplantation.

The prior allocation system grouped all LVAD complications into Status 1A, without distinguishing urgency. The revised system introduced risk stratification across defined complications. However, no study has evaluated the performance of this system as a rescue mechanism for LVAD patients experiencing complications. Prior investigations in the pulsatile LVAD era or in Europe focused primarily on post-transplant outcomes,^9–12^ introducing survival bias and failing to capture the overall impact of complications under allocation policies.

We therefore conducted a nationwide retrospective cohort study using the national database of the United States to examine outcomes among patients with HeartMate 3 LVADs listed under the revised UNOS allocation system and assess its performance as a rescue pathway and identifying areas for potential improvement.

## Methods

### Data Source

The United Network of Organ Sharing (UNOS) registry, a national transplant-specific database in the United States, was queried. This registry has prospectively collected patient-level data on the entire transplant population in the United States, including information on the donors, candidates, and recipients involved in every organ transplantation, since 1987. The data set used in this study was released on July 1, 2024.

This study utilized three components of the UNOS Standard Transplant and Analysis Research File: the Thoracic Main File, which contains patient information and clinical outcomes for each patient at the time of heart transplant registration and receipt; the Waitlist History File, which records all changes to the patient urgency status on the waiting list; and the Status Justification Form File, which catalogs qualifying reasons for urgency status changes. Data merging across these files as well as longitudinal analysis were permitted by utilizing unique identifiers assigned to each patient.

### Study Design and Participants

This study was a national registry retrospective cohort analysis investigating the outcomes of adults (≥18 years) with a HeartMate 3 LVAD who experienced a device-related complication resulting in either initial listing or status change between October 18, 2018, and June 30, 2023. UNOS defines eight complications as justification for status change: life-threatening ventricular arrhythmia, device malfunction/mechanical failure, device-related infection, mucosal bleeding, aortic insufficiency, thrombosis, right heart failure, and hemolysis. Due to small sample size (n=5), hemolysis was combined with thrombosis. The study period began at the UNOS policy allocation change in 2018, and ended one year before the data release date, based on the UNOS recommendations for data use.

This study was approved by the Program for Protection of Human Subjects at the Icahn School of Medicine at Mount Sinai. The approval included a waiver of informed consent.

### Study Endpoints

The primary outcome was all-cause mortality. Deaths recorded in the UNOS data set were ascertained through the Organ Procurement and Transplant Network. Secondary outcomes were heart transplantation and mortality on device.

### Statistical Analysis

Continuous variables were reported as means ± standard deviation and compared using the analysis of variance. Categorical variables were expressed as proportions and compared with Pearson’s chi-squared test.

We performed three analyses. First, post-complication outcomes were studied by estimating cumulative incidence of mortality on device, heart transplantation, and waiting on the list by using competing risk analysis with each event being mutually exclusive. Second, in a subgroup cohort of patients who underwent heart transplantation, post-transplant mortality risk was assessed using the Kaplan-Meier method. Risk-adjusted hazard ratios (HRs) for mortality were obtained by fitting multivariable Cox proportional hazard models adjusting for the following patient covariates: age, sex, race, blood type, body mass index, education level, heart failure etiology, functional status, prior cerebrovascular disease, diabetes, and dialysis. Third, post-complication mortality risk (regardless of transplant status) was assessed using the Kaplan–Meier method and Cox models with the aforementioned covariates.

All tests were 2-tailed: an alpha level of 0.05 was considered statistically significant. All statistical analyses were performed by using SAS version 9.4 (SAS Institute, Inc).

## Results

### Study Population

A total of 792 patients experienced an LVAD complication and had a corresponding status change during the study period. The patients’ baseline characteristics are shown in **Table 1**. The mean age was 52.0 ± 11.9 years and 81.3% were male.

**Table 1.**
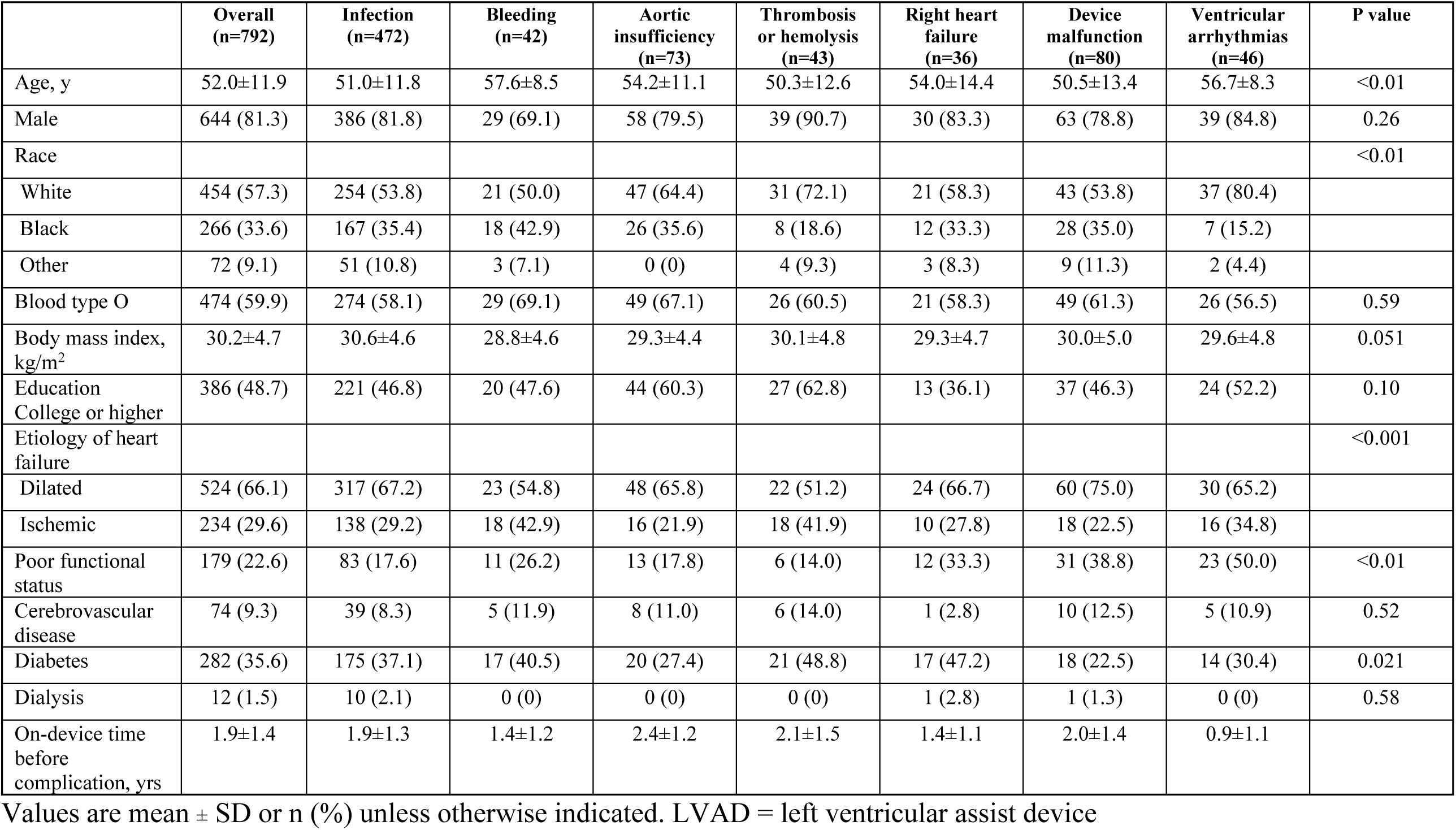
Baseline characteristics at LVAD complication occurrence based on complication types.

|  | Overall<br>(n=792) | Infection<br>(n=472) | Bleeding<br>(n=42) | Aortic<br>insufficiency<br>(n=73) | Thrombosis<br>or hemolysis<br>(n=43) | Right heart<br>failure<br>(n=36) | Device<br>malfunction<br>(n=80) | Ventricular<br>arrhythmias<br>(n=46) | P value |
| --- | --- | --- | --- | --- | --- | --- | --- | --- | --- |
| Age, y | 52.0±11.9 | 51.0±11.8 | 57.6±8.5 | 54.2±11.1 | 50.3±12.6 | 54.0±14.4 | 50.5±13.4 | 56.7±8.3 | <0.01 |
| Male | 644 (81.3) | 386 (81.8) | 29 (69.1) | 58 (79.5) | 39 (90.7) | 30 (83.3) | 63 (78.8) | 39 (84.8) | 0.26 |
| Race |  |  |  |  |  |  |  |  | <0.01 |
| White | 454 (57.3) | 254 (53.8) | 21 (50.0) | 47 (64.4) | 31 (72.1) | 21 (58.3) | 43 (53.8) | 37 (80.4) |  |
| Black | 266 (33.6) | 167 (35.4) | 18 (42.9) | 26 (35.6) | 8 (18.6) | 12 (33.3) | 28 (35.0) | 7 (15.2) |  |
| Other | 72 (9.1) | 51 (10.8) | 3 (7.1) | 0 (0) | 4 (9.3) | 3 (8.3) | 9 (11.3) | 2 (4.4) |  |
| Blood type O | 474 (59.9) | 274 (58.1) | 29 (69.1) | 49 (67.1) | 26 (60.5) | 21 (58.3) | 49 (61.3) | 26 (56.5) | 0.59 |
| Body mass index,<br>kg/m<sup>2</sup> | 30.2±4.7 | 30.6±4.6 | 28.8±4.6 | 29.3±4.4 | 30.1±4.8 | 29.3±4.7 | 30.0±5.0 | 29.6±4.8 | 0.051 |
| Education<br>College or higher | 386 (48.7) | 221 (46.8) | 20 (47.6) | 44 (60.3) | 27 (62.8) | 13 (36.1) | 37 (46.3) | 24 (52.2) | 0.10 |
| Etiology of heart<br>failure |  |  |  |  |  |  |  |  | <0.001 |
| Dilated | 524 (66.1) | 317 (67.2) | 23 (54.8) | 48 (65.8) | 22 (51.2) | 24 (66.7) | 60 (75.0) | 30 (65.2) |  |
| Ischemic | 234 (29.6) | 138 (29.2) | 18 (42.9) | 16 (21.9) | 18 (41.9) | 10 (27.8) | 18 (22.5) | 16 (34.8) |  |
| Poor functional<br>status | 179 (22.6) | 83 (17.6) | 11 (26.2) | 13 (17.8) | 6 (14.0) | 12 (33.3) | 31 (38.8) | 23 (50.0) | <0.01 |
| Cerebrovascular<br>disease | 74 (9.3) | 39 (8.3) | 5 (11.9) | 8 (11.0) | 6 (14.0) | 1 (2.8) | 10 (12.5) | 5 (10.9) | 0.52 |
| Diabetes | 282 (35.6) | 175 (37.1) | 17 (40.5) | 20 (27.4) | 21 (48.8) | 17 (47.2) | 18 (22.5) | 14 (30.4) | 0.021 |
| Dialysis | 12 (1.5) | 10 (2.1) | 0 (0) | 0 (0) | 0 (0) | 1 (2.8) | 1 (1.3) | 0 (0) | 0.58 |
| On-device time<br>before<br>complication, yrs | 1.9±1.4 | 1.9±1.3 | 1.4±1.2 | 2.4±1.2 | 2.1±1.5 | 1.4±1.1 | 2.0±1.4 | 0.9±1.1 |  |

Values are mean ± SD or n (%) unless otherwise indicated. LVAD = left ventricular assist device

Complications occurred a mean of 1.9 ± 1.4 years following LVAD implantation. The most common complication justifying status change was device infection (n=472, 59.6%), followed by device malfunction (n=80, 10.1%), aortic regurgitation (n=73, 9.2%), ventricular arrhythmias (n=46, 5.8%), thrombosis or hemolysis (n=43, 5.4%), bleeding (n=42, 5.3%), and right heart failure (n=36, 4.5%). Ventricular arrhythmias occurred the earliest after LVAD implantation at 0.9 ± 1.1 years, followed by right heart failure at 1.4 ± 1.1 years. Aortic insufficiency developed latest at 2.4 ± 1.2 years (**Table 1**).

Baseline characteristics across complication types differed significantly with respect to age, race, etiology of heart failure, baseline functional status, and diabetes (all p < 0.05). Other baseline characteristics were similar.

### Transplantation and Waitlist Mortality

Of the 792 patients, 573 (72.3%) patients underwent transplantation, with a mean time to transplantation of 128.5 ± 195.7 days. The 1-year cumulative incidence of transplantation was 71.5% (95% Confidence Interval [CI], 67.9-74.8%) (**Figure 1**). Device malfunction had the highest 1-year transplantation rate (92.5%, 95% CI, 81.9–97.0%) and a mean time to transplant of 38.3 ± 68.8 days. Ventricular arrhythmias had the fastest time to transplant (12.4 ± 16.8 days) and a 1-year transplantation rate of 84.8% (95% CI, 70.0–92.7%). The remaining complications had 1-year transplantation rates ranging from 53% to 83%, with mean times to transplant between 95 and 216 days. The 1-year cumulative incidence of waitlist mortality was 3.8% (95% CI, 2.5–5.4%) (**Figure 1**). Right heart failure carried the highest 1-year waitlist mortality (11.5%, 95% CI, 3.4–25.0%), followed by ventricular arrhythmias (8.7%, 95% CI, 2.6–19.7%) and device malfunction (4.2%, 95% CI, 1.0–11.3%). Other complications were associated with waitlist mortality rates of 2.4–3.9%.

**Figure 1.**
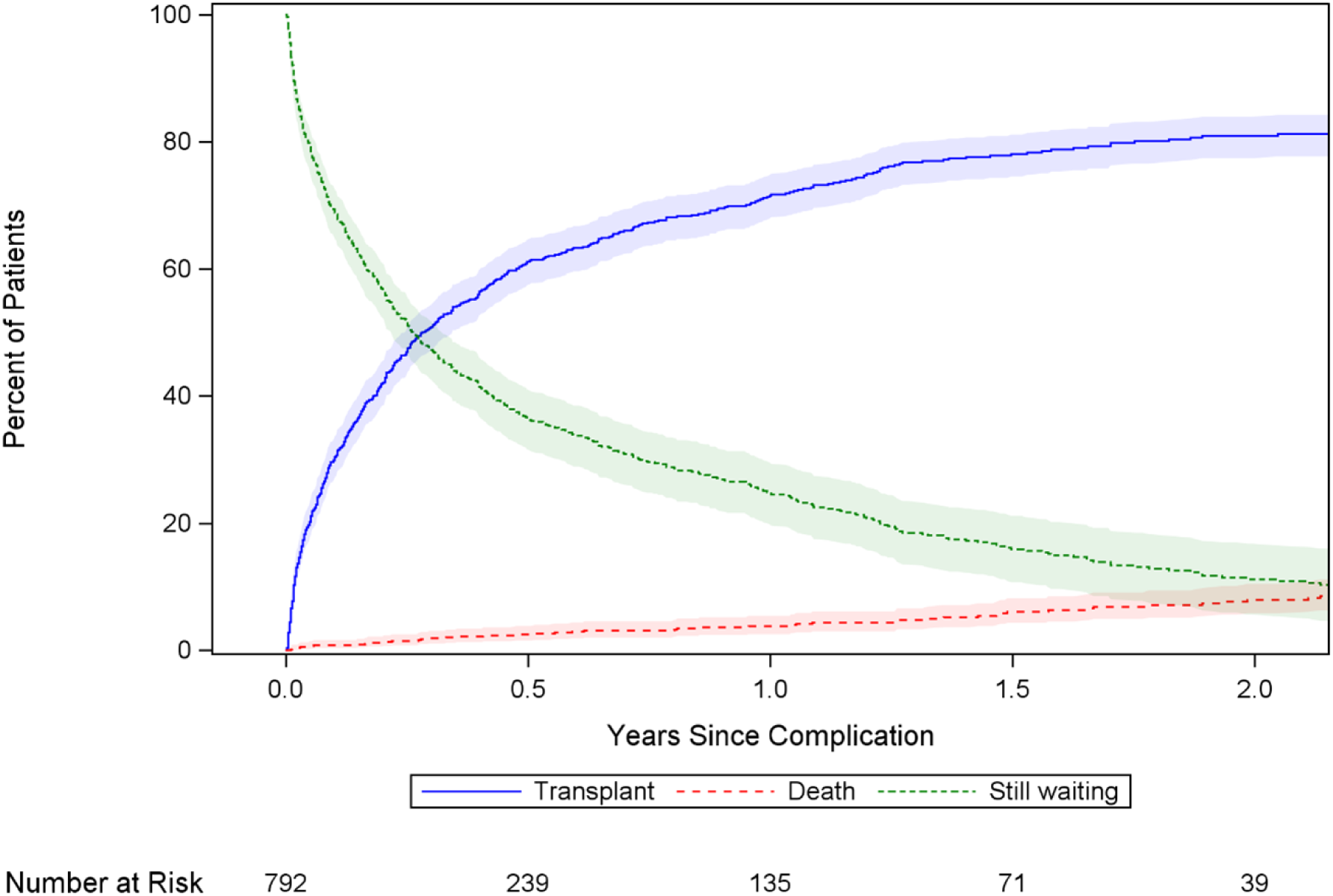
Competing Events After LVAD-Related Complication. Cumulative incidence of mutually exclusive events (mortality on device, heart transplantation, and remaining on the transplant waiting list) is plotted against time from the qualifying LVAD complication. Shaded areas indicate 95% confidence intervals. LVAD = left ventricular assist device.

### Post-Transplant and Overall Mortality

The 1-year post-transplant mortality was 12.5% (95% CI, 9.7–15.7%) (**Figure 2A**). Right heart failure had the highest 1-year post-transplant mortality (30.9%, 95% CI, 13.3–50.5%), though this was not significant after multivariable adjustment (HR 2.0, 95% CI, 0.9–4.9) (**Figure 2B**). Device malfunction (17.2%, 95% CI, 9.1–27.5%) and ventricular arrhythmias (15.3%, 95% CI, 5.4–30.0%) were associated with the next highest post-transplant mortality rates. Other complications ranged from 3.6–11.6%.

**Figure 2.**
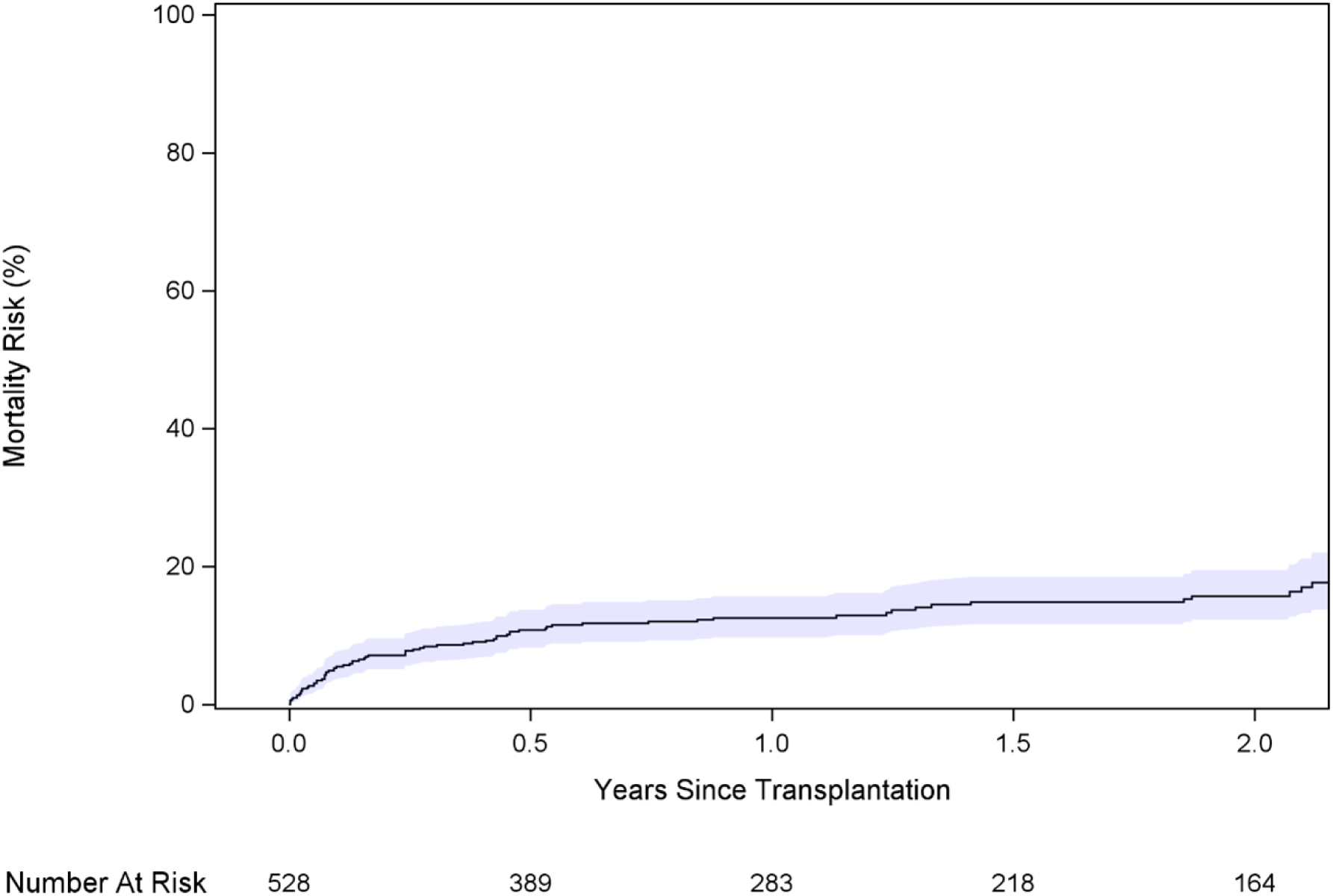
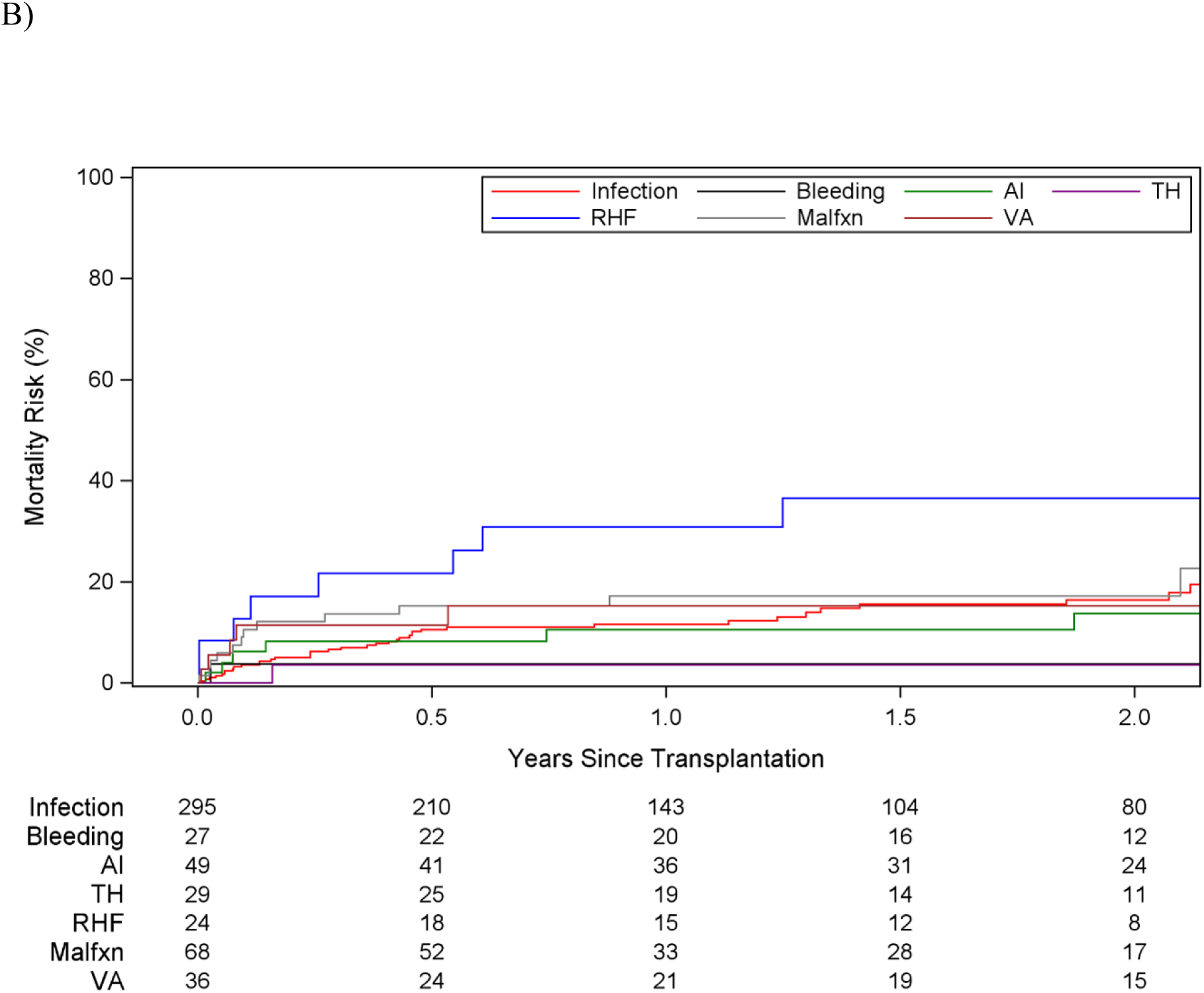
Mortality after Heart Transplantation. Post-transplant mortality risk is plotted against time from heart transplantation, among (A) the overall cohort and (B) stratified by complication type. Shaded areas indicate 95% confidence intervals. AI = aortic insufficiency; LVAD = left ventricular assist device; Malfxn = device malfunction; TH = thrombosis or hemolysis; RHF = right heart failure; VA = ventricular arrhythmia.

The 1-year overall mortality (including on-device and post-transplant deaths) was 12.3% (95% CI, 9.9–15.1%) (**Figure 3**). Right heart failure was associated with the highest overall mortality (34.1%, 95% CI, 18.2–50.8%) and was the only complication significantly associated with increased risk on multivariable analysis (HR 2.0, 95% CI, 1.1–3.8) (**Central Illustration**). Ventricular arrhythmias (22.6%, 95% CI, 10.9–37.9%) and device malfunction (17.6%, 95% CI, 9.9–27.2%) had the next highest overall mortality rates. Other complications ranged from 2.6–11.1%.

**Figure 3.**
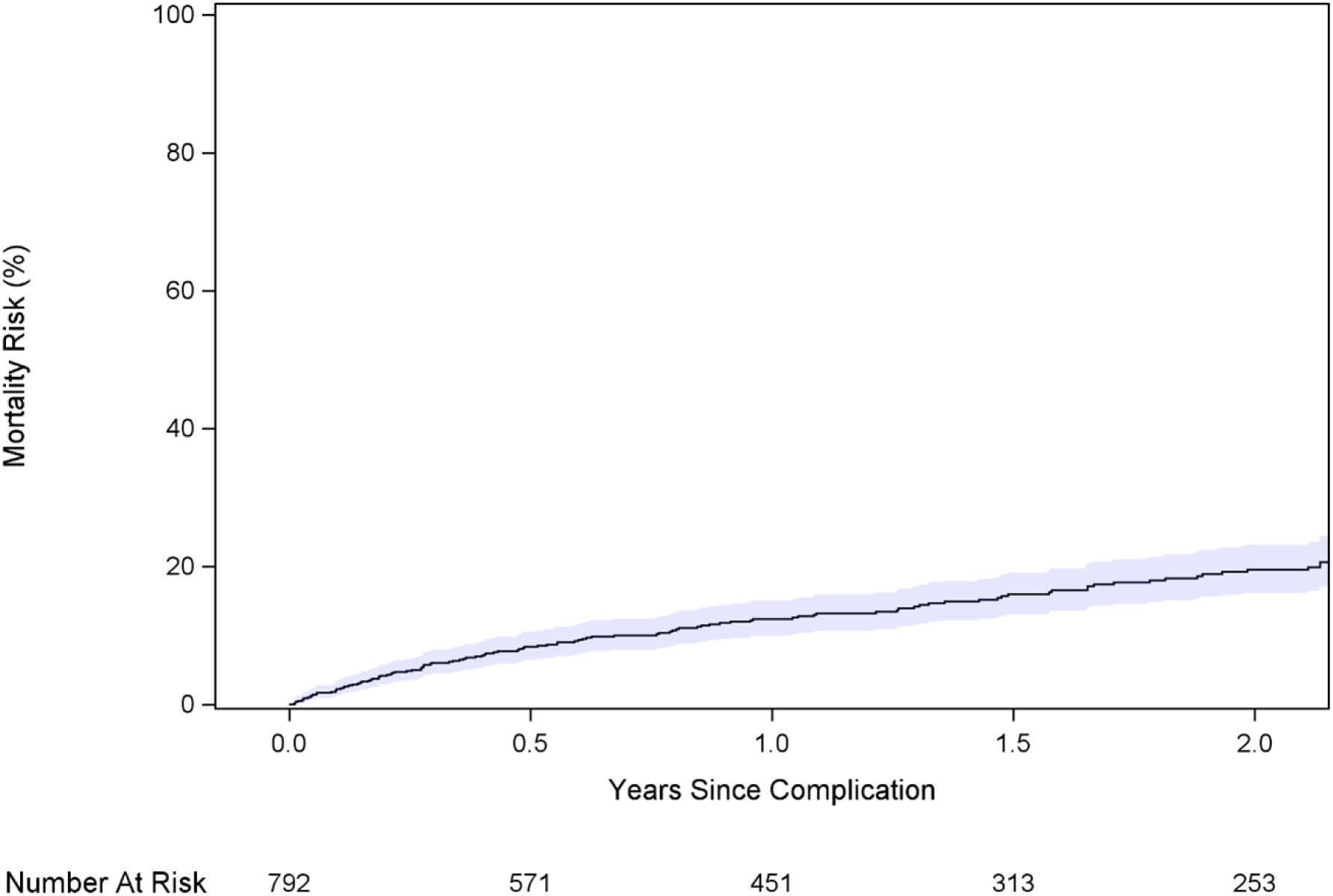
Mortality after LVAD-Related Complication. Mortality risk is plotted against time from qualifying LVAD-related complication among the overall cohort. Shaded areas indicate 95% confidence intervals. LVAD = left ventricular assist device.

## Discussion

In this nationwide investigation of HeartMate 3 LVAD patients experiencing complications under the revised UNOS allocation system, we found that the policy generally functions well as a rescue pathway, with high transplant rates and low waitlist mortality. However, right heart failure was a distinct exception, conferring twice the risk of mortality compared with other complications, with only two thirds of affected patients alive one year after onset.

In the prior allocation system, there were 5 categories of complications which allowed a status upgrade: thromboembolism, device-related infection, device malfunction, recurrent ventricular arrhythmia, and other complications. These complications all qualified for status 1A, the highest urgency tier. There was a concern that lack of discrimination among the complications may not give the most appropriate rescue pathway for each complication. The 2018 UNOS revision expanded and refined complication definitions, adding categories such as right ventricular failure, aortic valve insufficiency, mucosal bleeding, and hemolysis, and introduced risk-stratified urgency assignments (ventricular arrhythmia, status 1; device malfunction, status 2; remaining complications, status 3).

Our analysis showed that the new allocation system served well as a rescue mechanism for LVAD patients who experience LVAD related complications, by achieving a 70% transplant rate at 1 year and only 4% mortality on device at 1 year%, indicating that most patients who experienced complications were transplanted before developing irreversible organ dysfunction that would compromise post-transplant outcomes.

In terms of each complication, the new allocation system appears to have given adequate risk stratification overall. Ventricular arrhythmia (status 1) and device malfunction (status 2) had particularly high 1-year transplant rates (85% and 92%, respectively) and low waitlist mortality (9% and 4%, respectively). Among status 3 complications, right ventricular failure stood out: although 73% of these patients reached transplant by 1 year, waitlist mortality exceeded 10% and post-transplant 1-year mortality was 31%. In aggregate, only about two thirds of patients with right heart failure survived to 1 year, approximately double the risk observed with other complications.

Right ventricular failure after LVAD implantation presents unique clinical challenges.^13–15^ It commonly produces sustained hemodynamic compromise and end-organ dysfunction such as hepatic or renal impairment that may preexist device implantation and be less reversible despite LVAD support or transplantation. In that sense, the new allocation system correctly recognized right ventricular failure as a distinct category that deserves attention, although our findings suggest its current prioritization or timing of escalation may not fully address the excess mortality associated with this complication. The current UNOS definition requires at least moderate dysfunction (central venous pressure greater than 18 mmHg prior to medical intervention), and recognition depends on clinical judgment, which may delay status escalation. In our cohort, status escalation for right ventricular failure occurred a median of approximately 1.5 years after LVAD implantation. Greater awareness of the condition’s consequences and more liberal criteria for status upgrade could reduce time spent in a decompensated state and potentially improve outcomes.

Another plausible explanation for inferior outcomes is delayed access to transplant after status upgrade. Patients with right ventricular failure who ultimately underwent transplantation waited more than three months after upgrade, compared with an average of 50 days in the overall cohort. Shortening this interval could reduce cumulative end-organ damage and improve post-transplant survival.

Prior studies of LVAD complications, conducted in the pulsatile LVAD era or under European allocation systems, focused exclusively on post-transplant outcomes. ^9–12^ This approach introduced survival bias and failed to capture the full impact of complications. To our knowledge, this is the first study to evaluate outcomes from the time of status upgrade through transplantation in patients supported with HeartMate 3 under the revised UNOS system. This design allowed comprehensive assessment of the allocation system’s performance as a rescue mechanism in the contemporary LVAD era.

This study has several limitations. We analyzed only the first qualifying complication and could not assess cumulative or combined complications. The severity spectrum of the first qualifying complication was heterogeneous. Events met criteria for status upgrade but, at onset, were not severe enough to make patients transplant-ineligible; later progression to ineligibility was counted as a failed rescue (a hard endpoint in this study), so our findings may not capture the full range of clinical severity. We were also unable to capture outcomes for patients who obtained status upgrades via exception requests, which could introduce selection bias.

## Conclusions

The revised UNOS allocation system appears to provide LVAD patients experiencing complications an effective pathway to transplantation by achieving overall high transplant and low mortality rates. Right heart failure is an important exception with substantially worse outcomes with only two thirds of patients surviving to one year following this complication. These findings suggest that right heart failure should be considered for higher urgency status, and that improving early detection and shortening time to escalation may be necessary to improve survival.

## Data Availability

UNOS (the United Network for Organ Sharing) data; this database can be accessible for free for institutions which provides transplant upon request to UNOS.

https://unos.org/data/

## Acknowledgements

There are no acknowledgements for this study.

## Abbreviations and Acronyms

CI: confidence interval
HR: hazard ratios
LVAD: left ventricular assist device
REMATCH: Randomized Evaluation of Mechanical Assistance for the Treatment of Congestive Heart Failure
UNOS: United Network of Organ Sharing

---

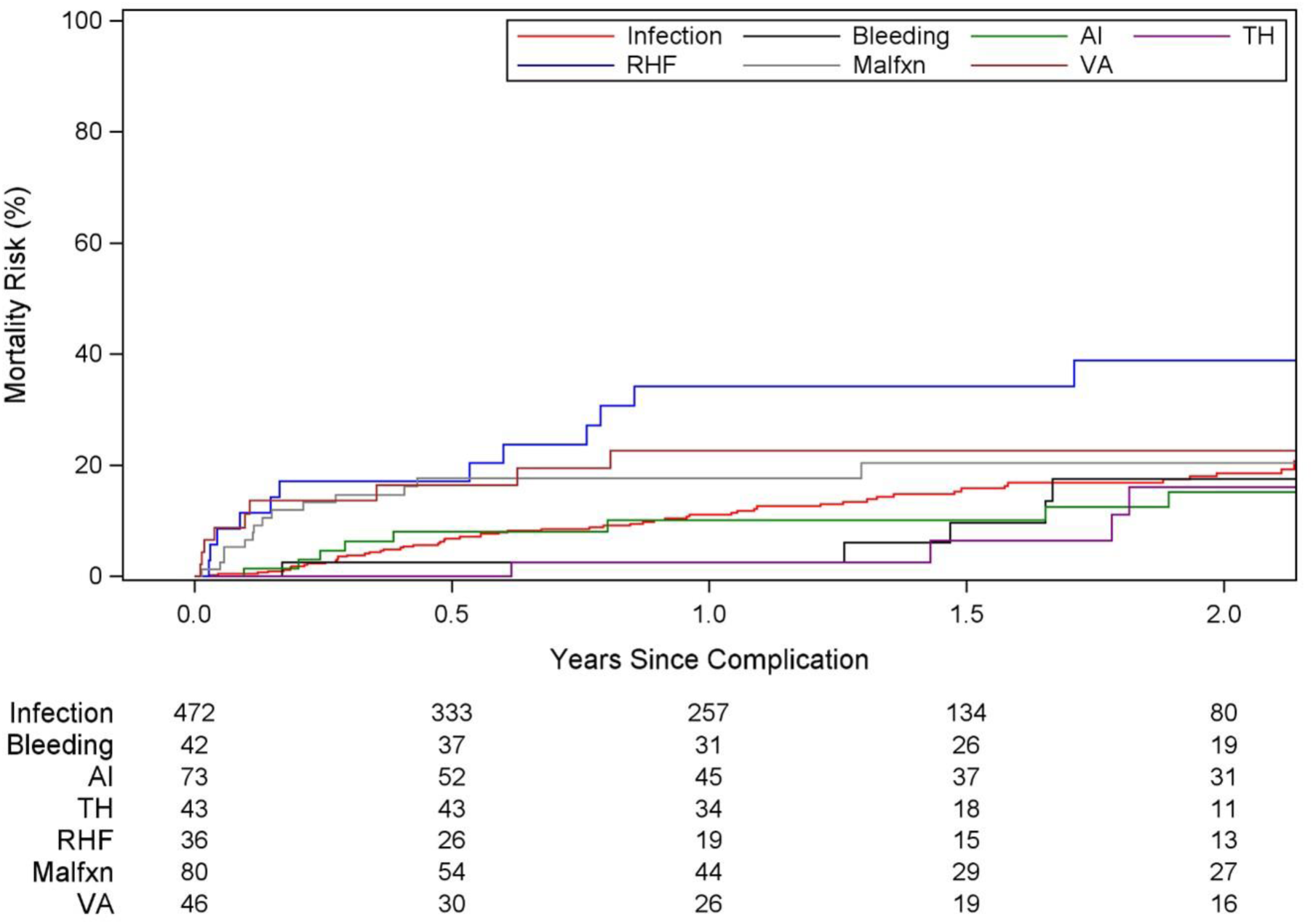
Central Illustration. Mortality after LVAD-Related Complication, Stratified by Type. Mortality risk plotted against time from LVAD-related complication, stratified by complication types. Right heart failure was associated with a 1-year mortality of 34.1% (95% CI, 18.2-50.8%) and was the only complication associated with a significant increase in 1-year mortality with a hazard ratio of 2.0 (95% CI, 1.1-3.8). AI = aortic insufficiency; LVAD = left ventricular assist device; Malfxn = device malfunction; TH = thrombosis or hemolysis; RHF = right heart failure; VA = ventricular arrhythmia.

